# Prevalence and monitoring utility of glycated albumin among diabetic patients attending clinic in tertiary hospitals in Dodoma, Tanzania: A cross-sectional study protocol

**DOI:** 10.1101/2023.07.19.23292868

**Authors:** George Gabriel Mkumbi, Matobogolo Boaz

## Abstract

Diabetes mellitus is a serious public health concern, with third world nations accounting for 80% of the new cases. Tanzania has a high diabetes burden, with rising prevalence, complications, and death, as well as life-threatening impairments. Diabetic diagnosis and prognosis are currently based on two tests: plasma glucose and glycated hemoglobin (HbA1c). However, other markers of glucose homeostasis, such as fructosamine and glycated albumin (GA), may be seen as an appealing alternative, especially in patients whose HbA1c test is skewed or incorrect. GA appears to have a higher overall diagnostic efficiency than fructosamine in a variety of clinical contexts. Further research is needed to determine whether GA can complement or replace traditional glycemic control measurements like HbA1c, as GA may aid in the therapeutic management of diabetic individuals whose HbA1c levels are unreliable.

**Method:** A hospital-based cross-sectional analytical study design will be conducted among diabetic patients attending the diabetic clinics of the Dodoma Regional Referral Hospital and Benjamin Mkapa Hospital from 1^st^ August to 30^th^ October, 2023.All patients with a diagnosis of diabetes mellitus for more than 6 months on medication will be screened for eligibility. Informed consent, history and clinical examination will be obtained in all the patients. All patients will be subjected to voluntary blood sample collection and blood samples obtained will be sent for RBG and HbA1c. Simultaneously the Glycated Albumin levels will be obtained from the same blood samples collected. Standard glycemic status of all patients will be defined as per HbA1c. A level greater than 7% will be considered as a poor indicator, hence poor control in patients with more than six months of treatment. A questionnaire containing both open and closed ended questions will also be used in recording the patient’s response. Analysis including both descriptive and inferential statistics will be computed with SPSS version 28.0. and a predictor variable P<0.05 will be considered as statistically significant.

## Introduction

Diabetes mellitus has been one of the twenty-first century’s most serious healthcare challenges [1]. The disease is spreading at an alarming rate throughout the world, with developing nations accounting for 80% of the new cases [2]. In 2015 it was reported to affect 8.8 percent (415 million) of adults worldwide, and it is expected that 652 million individuals (10.4 percent) would have diabetes by 2040 [3]. Tanzania, like the rest of Sub-Saharan Africa, has a high diabetes burden, with rising prevalence (14.8%), complications, and death, as well as life-threatening impairments [4,5].

In order to limit the risk of diabetes complications, diabetic patients must be properly diagnosed and monitored [6]. Currently, diabetes diagnosis and prognosis are mostly dependent on two tests: serum/blood glucose and glycosylated hemoglobin (HbA1c) [7]. These metrics, however, are not failsafe, and their therapeutic utility is influenced by a variety of clinical and analytical parameters [8]. Other glucose homeostasis markers, like as in fructosamine and glycated albumin (GA), may be viewed as a desirable alternative, particularly in individuals whose HbA1c test results are inaccurate. Individuals with hemoglobinopathies and kidney illness, for instance, exhibit rapid alterations in glucose homeostasis and greater glycemic excursions [9,10]. GA appears to have a higher overall diagnostic efficiency than fructosamine in a variety of clinical contexts, according to existing information [11].

GA is a glycemic control indicator that has been examined as a substitute for HbA1c in people with diabetes mellitus. GA is a far more dependable glycemic variability indicator than HbA1c. Accumulating evidence suggests that GA is indeed a precise diagnostic test that is directly related to microvascular complications in diabetes [8]. Additionally, it is suitable for individuals’ undergoing hemodialysis and its levels are unaffected by anaemia or haemolytic processes. GA is preferable to fructosamine because it is not impacted differently by different serum proteins. The catalytic technology used to analyze it is easy to deploy and quick to execute, in addition to being highly effective analytically and possessing a greater degree of standardization [12]. In clinical situations where HbA1c values are erroneously altered, GA testing may provide a reliable result for monitoring DM, according to a recent study. This is owning to the fact that the physiological mechanisms by which these two glycated proteins are produced; guarantees that GA outperforms HbA1c in terms of the assessment of glucose homeostasis in the absence of confounding factors [8].

Additional advantages of glycated albumin over glycosylated hemoglobin include decreased reagent costs and the possibility to automate glycated albumin measurement on numerous conventional laboratory instruments [13]. Although additional research is required to evaluate whether GA could augment or even replace conventional diabetic markers such as HbA1c; GA may assist in the diabetic care of patients with inaccurate HbA1c readings [14]. As a final regard to the fore mentioned benefits of GA; an international consensus on clinical usage is required to ensure its inclusion in routine clinical laboratory workup, hence improving future monitoring and management of DM patients. For this to happen a lot of similar research like the one proposed, are required.

## Materials and Methods

### Study aims

1. To determine the prevalence of poor glycemic control among diabetic patients attending clinics in Dodoma Tertiary hospitals.
2. To determine the validity of serum glycated albumin as an index of glycemic control in diabetic patients attending clinics in Dodoma Tertiary hospitals.

### Study design

A cross sectional study design will be conducted at the Benjamin Mkapa Hospital and the Dodoma Regional Referral Hospital in Dodoma, Tanzania; for a period of three months.

### Study setting

The medical centers at both BMH and DRRH will serve as the study sites. Dodoma, the capital city of Tanzania, is home to several medical facilities. Dodoma Regional Referral Hospital (DRRH) and Benjamin Mkapa Hospital (BMH) are both referral hospitals for the Dodoma region, the central zone and surrounding areas respectively. In addition, the University of Dodoma uses both hospitals as teaching centers (UDOM). Health care for those living in the central zone is facilitated in part by these facilities. The population of the Dodoma region was 2.49 million as per the 2012 Population and Housing Census.

BMH and DRRH have a capacity of 500 and 420 beds respectively. Each month, approximately 1400 patients with medical related diagnosis are seen in the medical department at BMH, with fifteen to twenty (15-20) of these patients being diagnosed with diabetes mellitus seen everyday. Additionally, 480 patients with diabetes are documented at DRRH each month. Thus, a total of 10-15 patients with diabetes mellitus are typically seen per clinic session and a total of 30-35 patients per session from both health facilities combined (Unpublished data).

### Sample size estimation

Following the Leslie-Kish formula [15,16], the minimum sample size required to include a patient in the study will be determined as below;

Where:

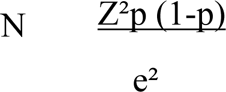

N = Minimal sample size

Z = The value from the normal distribution scale that indicates a degree of significance (1.96 for 95% confidence level).

p = 84.3 % is the rate of uncontrolled blood sugar among diabetic patients who visit the outpatient clinic [17]

e = margin of error (usually 5%) = 0.05

So;

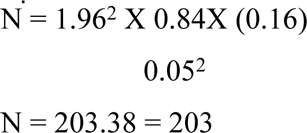

***<u>Therefore, the total sample size will be 203 participants</u>***

### Inclusion criteria

1. Patients with at least 18 years of age.
2. Patients who will accept participation in the study through signing an informed consent.
3. Patients who are on OAD, insulin, or a combination of the two for at least six months.

### Exclusion criteria

1. Patients who have received blood transfusions during the last three months.
2. Patients who are at risk of receiving or donating blood / blood products over the course of the study.
3. Patients who are receiving erythropoietin.
4. Patients who are receiving Iron supplements.

### Participants characteristics

The study participants will be adults (18 years and older) who have been diagnosed with diabetes mellitus and are attending the diabetic clinics at Benjamin Mkapa Hospital and Dodoma Regional Referral Hospital during the study period.

### Process

#### Participant interview

A participant who provided informed consent will be interviewed using a structured questionnaire that collects demographic data such as age, gender, and level of education. In addition, a minimum of two contacts; one of the patients and the second one of the next of kin will be recorded. As part of social demographic data alcohol consumption history (defined as alcohol consumption within the past 12 months), with an emphasis on duration and frequency will be collected [18,19]. In addition to the above also duration and number of cigarettes in the number of packs per day, the history of current smoking, defined as smoking within the past year, will be obtained [20]. A history of hypertension (defined as a history of hypertension or the use of antihypertensive medications) and diabetes type (defined as a history of diabetes or the use of diabetic medications) will be documented [21].

### Clinical examination

A blood pressure (BP) reading will be taken using an automated digital machine of AD Medical Inc. brand, keeping with the 2018 AHA/ACC Hypertension guideline for standard measurement of BP [22]. Hypertension will be defined as BP ≥140/90 mmHg, in a patient with a history of hypertension, or a patient on antihypertensive medications [23]. The radial pulse will be measured using a finger pulse oximeter model FL–100.

The measurements for all of the patients will be done by two operators; Operator 1 will listen to the heart sounds with a stethoscope while operator 2 will palpate the radial pulse while the patient is seated. When both operators are ready, a timer will be utilized to begin counting at the same time. The technique will be repeated, and the heart rate will be recorded after 1 minute. To eliminate bias, the two operators will switch roles in different patients. The difference between the rates counted by the two operators at the end of one minute will be used to determine the apex-pulse deficit [24]. A deficit of ten or more will be considered indicative of atrial fibrillation [25]. In addition, each operator will assign a regular or irregular rhythm to the beat.

### Sampling technique

In this study, the consecutive sampling method will be used to recruit the participants. This will enable the researcher to target and track the available patients as they visit the clinic. Therefore, the sampling technique will allow the researcher to get in contact with any patient meeting the criteria.

### Data collection methods

This study will collect its data through the use of a questionnaire provided by the researchers, which will consist of both open-ended and closed-ended questions. The data collection process will begin in August 2023 and will go until October 2023. Three research assistants will be involved and trained by the principal investigator to guarantee that the questionnaires are filled out correctly and that no research misconduct occurs during data collecting. Two certified nurses and one laboratory technician will serve as researcher assistants. Each assistant interviewer will be trained to ask each participant the same pertinent questions listed in the questionnaire in order to minimize researcher’s bias during data collection. Respondents will be screened before the start of the clinic, and the questionnaires will be completed after informants have received service.

Participants who consent to participate in the study will be recruited, and their biodata will be collected using a systematic, pre-tested questionnaire administered by an interviewer. Their blood pressure will be gauged using an Accoson mercury sphygmomanometer. The systolic and diastolic blood pressures will be estimated using Korotkoff’s sounds, namely the first and fifth phases. Each individual’s kilogram weight will be measured using an RGZ-120 Health weighing scale and rounded to the nearest decimal place. Participants will be requested to stand firmly on a scale while facing the investigator. They will be instructed to remove their shoes, wear minimal clothing, and place their things on a nearby table. Next, we’ll check the scale’s readout to determine their body mass. A stadiometer will be used to ascertain each person’s height (RGZ-120 Health scale). Participants will be asked to take off their shoes and hats and stand with their backs to the researcher. Once the scale will be adjusted so that it touches the top of the participant’s head; and a direct reading will be taken and converted to meters to within one decimal place. Their body mass index (BMI) will be determined by dividing their weight in kilograms by the square of their height in meters (kg/m2).

### Laboratory procedure

Following completion of the questionnaire, respondents will be asked to consent to a venepuncture, in which three millilitres of blood will be drawn on either side of the cubital fossa using a 5cc syringe and placed in a red and lavender top vacutainer tube. The blood sample for both study blood collection centres will be transported to the authorized Benjamin Mkapa Hospital (BMH) laboratory in a refrigerated box with an ice pack for planned tests. Blood samples will be processed according to the BMH laboratory’s Standard Operating Procedures for each test.

### Data collection tools

This study will use standardized, structured surveys with closed and open-ended questions as its data gathering instrument. The questionnaire will consist of three parts: the first will assess the patient’s sociodemographic characteristics; the second will evaluate the patient’s clinical characteristics; and the third will include laboratory results and bio data like the patient’s glycated haemoglobin and haemoglobin A1c levels, body mass index, blood pressure, fasting blood glucose, and random blood glucose levels.

If respondents choose to fill out the surveys on their own, we will provide them with a Swahili translation. Enzymatic determination of GA will be performed using a liquid reagent (Lucica GA-L® Asahi Kasai Pharma Co., Tokyo, Japan) that consists of ketoamine oxidase and an albumin-specific proteinase, and the results will be evaluated using a Cobas 6000 series HPLC analyser. Automated high-performance liquid chromatography from the National Glycated Haemoglobin Standard Program, and the International Federation of Clinical Chemistry will be used to calculate the HbA1c (percent) value.

### Validity of the data collection tools

Validity will be defined as the degree to which tools or tests accurately measure what should be measured [26]. The standardized equipment and structured questions used in this study will be re-evaluated to ensure that they are appropriate for the local situation and adequately assess the required variables. The draft version of the structured instrument and questions for measuring the variables will be provided to specialists from the University of Dodoma’s School of Medicine and Dentistry and to the supervisors. Modifications will be made in consultation with specialists and the supervisor. During the pilot project, data gathering tools will be pre-tested.

### Reliability

The term “reliability” will refer to the state of being accurate or precise, as well as the consistency of the data gathered during the investigation [26]. The glycated haemoglobin test is considered the gold standard for monitoring glucose levels, and its sensitivity and specificity will serve as the basis for this study’s reliability (HbA1c). Additionally, structured questionnaires will be used to elicit information from informants. To guarantee the data’s reliability, the study’s tool will be pre-tested to determine its correctness and ability to produce the desired results.

### Pilot study

A pilot study defined as a smaller-scale version of a larger-scale study that is conducted to improve and clarify aspects of the study’s methodology, such as the tools and processes to be utilized in data gathering will be performed [27]. The purpose of a pilot study is to determine whether or not the full-scale investigation is feasible and to reveal study strengths and flaws. In this study, a field testing will be performed with 10% of the minimal sample size at DRRH in Mid-July. The pilot study will specifically test for adequacy, feasibility and estimation of the time required for filling questionnaires so as to identify the problems that may occur during research work. The information obtained from pilot study is hoped to bring facilitation in conducting the proposed study.

### Measurements of the variables

#### 1. Dependent variables

The levels of glycated albumin GA and glycated haemoglobin HbA1c will be evaluated in the study population. The two glycated proteins used are biomarkers for evaluating glycemic control in diabetic patients and will serve as the study’s dependent variables. Normative values will be used to assess these variables.

#### 2. Independent variables

##### I. Demographic variables

We will use demographic variables such as respondent age, gender, marital status, education, area of residence, and method of healthcare finance to better understand the needs of our informants. Nominal indicators will be used to classify participants according to sex, healthcare coverage, marital status, and geographic location. We will use accurate age and educational background in ordinal scales. All of the demographic information for this study will be gathered by questionnaires.

##### II. The patient’s glycaemic control

The glycation process and, by extension, levels of advanced end products of glycation, are directly influenced by factors such as the patient’s physiological and pathological profile, including pregnancy, racial heritage (primarily black/African origin), comorbidities (Anaemia, CKD, Hemoglobinopathies), and dialysis treatment. As a result, these are our dependent variables and through which numerous biomarkers fore mentioned, are affected.

### Data analysis plan

After data entry into the master coding sheet is complete, it will be analysed using SPSS version 28.0. The data will be cleaned and completeness checked by calculating the frequency of all variables. Descriptive statistics will be utilized to examine the demographics of the respondents. The percentage and median ages will be published, and the percentages of each gender, place of residence, degree of education, and marital status will also be shown. The percentage of those with poor glycemic control will be calculated by dividing the number of respondents with increased HbA1c values by the total number of respondents who had the test.

The findings will be shown in the form of a count (and percentage), a mean (and standard deviation), and a median (25th-75th percentiles). Comparisons of baseline characteristics between glycemic-control subgroups will be made using chi-square, analysis of variance, and Kruskal-Wallis tests. The correlation between two continuous measures will be estimated using the covariance estimate of the multivariate t distribution. This will provide some resistance to extremes without an excessively high breaking threshold. Using a single dependent variable, two correlation coefficients will be calculated and the significance of their difference will be tested using the Williams’ test. Segmented regression analysis will be used to examine the ties between indicators of glucose homeostasis.

For possible piecewise linear relations, this method of regression generates separate regression coefficients. The split point between two segmented relations will be calculated using the outcomes of the Davies’ test for a non-zero difference in slope between variables. GA % and HbA1c’s ability to predict the existence of impaired glucose tolerance will be assessed and compared using the area under the receiver operating characteristic curve (AUC, C-statistic). Youden’s J-point will be used to establish diagnostic cut-offs for the different types of glucose intolerance. The Kappa statistic will be used to measure the degree of agreement between markers at these arbitrary cut-off points, and the 95% CI will be determined through the use of 2000 replicates with a stratified bootstrap and percentile approaches. These cut-offs will be used to assess the performance of GA percent and HbA1c using the following performance measures (with a 95% confidence interval): sensitivity, specificity, Youden’s Index, positive predictive value (PPV), negative predictive value (NPV), accuracy, diagnostic odd ratio (DOR), number needed to diagnose (NND), likelihood ratio of a positive test (LR+), and likelihood ratio of a negative test (LR-). Assuming parallel testing, we will additionally assess the diagnostic utility of combining GA % and HbA1c. If the p-value is less than 0.05 on two-sided testing, then the result is considered statistically significant with a 95% CI. Cronbach’s alpha will be used to check the data’s reliability. The study will report its findings in accordance with the Standards for the Reporting of Diagnostic Accuracy Studies (STARD).

### Ethical issues

Ethical considerations will be taken throughout this project, from participant recruitment to data collection through analysis to dissemination of results. This will help make sure researchers are abiding by all local, regional, and national guidelines. The participants will be briefed about the nature and goals of the study. Participants who sign an informed consent form and otherwise meet the study’s inclusion criteria will be enrolled.

Participants will be able to revoke their permission at any moment during the course of the study. Participants who withdraw their consent at any point during the study period will get the same level of treatment as those who choose not to participate. The participants’ veins will be pricked and a total of 3 millilitres of blood will be drawn into two separate vacutainers for use in the lab.

The respondents’ names will be removed from the questionnaires and a coding system will be used to represent their information throughout the data processing process. All responses and study results will be kept strictly confidential and utilized for academic analysis. Also, in compliance with the standards set by the University of Dodoma, the results of this study will be disclosed for the sole purpose of formulating an action plan. In order to better the health of study subjects, the researcher will alert the proper authorities, such as the Tanzanian Ministry of Health and the management of the chosen institutions, to follow up with those respondents who have laboratory findings requiring quick care.

The UDOM institutional research review ethics committee has provided its blessing to the PI’s request for ethical approval through a letter with Ref No. MA.84/261/02/9. This confirmation letter from the University of Dodoma was presented to the regional administrative secretary for the Dodoma region and the hospital administration, and permission to collect the data was granted in both hospitals though letter With Ref No. DB.122/467/01G/29 And GA.244/292/01/’E’/45 respectively.

### Study timeline

The duration of this study will be three months, from 1^st^ August to 30^th^ October. Two months of data collection and one month of data analysis and write up.

## Discussion

Glycemic monitoring is an important part of diabetes therapy, and glycated albumin (GA) and glycated haemoglobin (HbA1c) are two typical indicators [28]. Although both are helpful in tracking how well diabetes is being managed, they are measured and used clinically in different ways. whereas HbA1c represents glucose levels on average over the previous two to three months, which is the usual lifespan of a red blood cell. Haemoglobin beta chain glycosylation occurs when glucose covalently bonds to the amino acid valine at the N-terminal end of the beta chain [29]. Several guidelines for the diagnosis and management of diabetes advocate measuring HbA1c as a benchmark [30]. GA, on the other hand, is generated when glucose binds to albumin without the help of an enzyme, and hence represents glycemic control during the previous two to three weeks [31]. GA forms at a much quicker rate than HbA1c [32].

Both HbA1c and GA can be used as indicators of glycemic control, but each has its own unique clinical applications [33]. There is a high correlation between HbA1c levels and the risk of microvascular and macrovascular problems in diabetes, and HbA1c has been widely utilized as a marker of glycemic control in the disease [29]. The American Diabetes Association (ADA) recommends that most individuals with diabetes aim for a HbA1c of 7%, with a target of 6.5% for some patients [34,35]. However, there are specific clinical contexts where HbA1c cannot be reliably used. HbA1c readings may be misleading, for instance, in those with hemoglobinopathies or anaemia. Haemolysis, blood transfusion, and other disorders that impact red blood cell turnover can also affect HbA1c levels [30,36,37]. However, liver illness, pregnancy, and other diseases that modify albumin turnover may have an effect on GA [37]. While HbA1c can be impacted by factors like erythrocyte turnover and drugs that shorten or lengthen the lifespan of erythrocytes, GA is unaffected by these things and its measurement is less prone to inconsistency (6). Patients with chronic kidney disease (CKD) may also benefit from GA [38,39]. Falsely low HbA1c values in CKD patients may be caused by variables such erythropoietin insufficiency, anaemia, and erythrocyte fragmentation. On the other hand, GA is not influenced by these factors and may be a more reliable indicator of glycemic control in CKD patients [37].

Several studies have shown that GA levels are linked to HbA1c levels and the risk of microvascular and macrovascular problems in diabetes [40]. One study indicated that GA levels were substantially related to the danger of diabetic eye disease and kidney disease [41]. Additional research has linked elevated GA levels to type 2 diabetics’ risk of developing and experiencing severe coronary artery disease [37,40]. A recent study also suggested that GA, rather than HbA1c, may be a more accurate predictor of cardiovascular events in those with type 2 diabetes [42].

When comparing glycated albumin to HbA1c for purposes of diabetes control, a cross-sectional study has various limitations. Uncertainty over timing, potential for bias in selection or measurement, and restricted applicability are all identified as potential limitations. Care has been taken to address these drawbacks multiple ways as; Selection bias will be reduced with careful sampling, and measurement bias can be corrected with tried-and-true methods detailed earlier. The findings are usually more broadly applicable if they are based on a diverse and representative study population, hence two centers that serve a diverse group of diabetic patients that are coming from the nation’s capital were chosen. These methods aim to improve the trustworthiness and validity of the findings.

In conclusion, glycated hemoglobin (HbA1c) is the current gold standard marker for monitoring glycemic control, which is critical in the management of diabetes. GA may offer some benefits over HbA1c in certain therapeutic settings, despite HbA1c being a well-known marker with standardized measurement and documented clinical importance. With a shorter half-life and more consistent measurement, as well as immunity to confounding factors including erythrocyte turnover and blood transfusion, GA shows promise as a helpful marker of glycemic control in individuals with diabetes. This is especially true for people whose glycemic control is less stable or who have chronic renal disease. Despite GA not being as frequently investigated as HbA1c, it has been linked in a number of studies to microvascular and macrovascular problems in diabetes. Therefore, it’s the goal of the current study to show the true therapeutic utility of GA and its applicability to our settings.

## Data Availability

No datasets were generated or analysed during the current study. All relevant data from this study will be made available upon study completion.

## Dissemination of the findings

The final results will be submitted to the University of Dodoma library, the study areas (Dodoma regional referral hospital, and the Benjamin Mkapa Hospital) and a manuscript will be submitted in different peer-review journals for publication.

## Authors’ contributions

Conceptualization, Data curation, Formal analysis, Investigation, Methodology, Resources, Writing – original draft, Writing – review & editing: George Gabriel Mkumbi.

Conceptualization, Methodology, Project administration, Resources, Supervision, Writing – review & editing: Matobogolo Boaz.

## Acknowledgements

The authors would like to thank the participants and staff of Dodoma regional hospital and Benjamin Mkapa hospital for donating their time to this project.

## Appendix

Aim 1
| **Variable** | **Method of measurement** | **Operational definition** | **Level of measurement** |
| --- | --- | --- | --- |
| Glycemic Control | blood sample | HbA1c level < 7% | Dichotomous |

Aim 2
| **Variable** | **Method of measurement** | **Operational definition** | **Level of measurement** |
| --- | --- | --- | --- |
| Gold standard Test for Glycemic Control<br>(Glycated Haemoglobin **HbA1c**) | blood sample | HbA1c level < 7% | Dichotomous |
| Experimental Test for Glycemic Control<br>(Glycated Albumin **GA**) | blood sample | GA<22% | Dichotomous |

